# Measures of socioeconomic advantage are not independent predictors of support for healthcare AI: subgroup analysis of a national Australian survey

**DOI:** 10.1101/2023.01.30.23285209

**Authors:** Emma Frost, Pauline O’Shaughnessy, David Steel, Annette Braunack-Mayer, Yves Saint James Aquino, Stacy M. Carter

## Abstract

Applications of AI (artificial intelligence) have the potential to improve aspects of healthcare. However, studies have shown that healthcare AI algorithms also have the potential to perpetuate existing inequities in healthcare, performing less effectively for marginalised populations. Studies on public attitudes toward AI outside of the healthcare field have tended to show higher levels of support for AI amongst socioeconomically advantaged groups that are less likely to be sufferers of algorithmic harms. We aimed to examine the sociodemographic predictors of support for scenarios related to healthcare AI.

The AVA-AI survey was conducted in March 2020 to assess Australians’ attitudes toward artificial intelligence in healthcare. An innovative weighting methodology involved weighting a non-probability web-based panel against results from a shorter omnibus survey distributed to a representative sample of Australians. We used multinomial logistic regression to examine the relationship between support for AI and a suite of sociodemographic variables in various healthcare scenarios.

Where support for AI was predicted by measures of socioeconomic advantage such as education, household income, and SEIFA index, the same variables were not predictors of support for the scenarios presented. Variables associated with support for healthcare AI across all three scenarios included being male, having computer science or programming experience, and being aged between 18 and 34 years. Other Australian studies suggest that this group have a higher level of perceived familiarity with AI. Our findings suggest that while support for AI in general is predicted by indicators of social advantage, these same indicators do not predict support for healthcare AI.

**WHAT IS ALREADY KNOWN ON THIS TOPIC:** Artificial intelligence has the potential to perpetuate existing biases in healthcare datasets, which may be more harmful for marginalised populations. Support for the development of artificial intelligence tends to be higher amongst more socioeconomically privileged groups.

**WHAT THIS STUDY ADDS:** Whilst general support for the development of artificial intelligence was higher amongst socioeconomically privileged groups, support for the development of healthcare artificial intelligence was not. Groups that were more likely to support healthcare artificial intelligence were males, those with computer science experience, and younger people.

**HOW THIS STUDY MIGHT AFFECT RESEARCH, PRACTICE, OR POLICY:** Healthcare artificial intelligence is becoming more relevant for the public as new applications are developed and implemented. Understanding how public attitudes differ amongst sociodemographic subgroups is important for future governance of healthcare AI.

## BACKGROUND

There are currently many applications for healthcare artificial intelligence (HCAI) in various stages of development and implementation [1]. Defined as technologies that allow computer programs to perform tasks and solve problems without explicit human guidance [2], in healthcare artificial intelligence-based systems employ algorithms to complete tasks typically performed by health professionals. Algorithms have been trained to read electrocardiograms [3], detect skin cancer from smartphone images [4], and predict people’s risk of disease using large scale national datasets [5] with ostensibly comparable accuracy to current approaches.

Whilst these technologies have the potential to improve aspects of healthcare, they also have the potential to cause harm to patients [6]. Algorithmic harms are exacerbated in already marginalised populations [7,8], as the causes and effects of historical structural disadvantage are embedded in healthcare datasets, and training sets often exclude marginalised groups. Obermeyer et al. [9] audited an algorithm used in the US for determining whether patients should be referred to high risk care, and found that patients who identified as Black were less likely to be flagged by the algorithm as needing high-risk care, despite having more comorbidities than non-Black-identifying counterparts. Similarly, Seyyed-Kalantari et al. [8], using data from the US, found that women, people aged under 20, those with lower socioeconomic status, and Black or Hispanic-identifying people were less likely to be diagnosed correctly by a chest radiograph algorithm. Factors preventing marginalised groups from accessing care in the past exist implicitly in many healthcare datasets, and algorithms trained on these datasets perpetuate these inequities [9].

Surveys examining public attitudes towards AI have found that certain sociodemographic characteristics are associated with higher levels of support for AI. Zhang and DaFoe [10] in a survey in the US, found that younger people, males, those with computer science experience, and those with a high annual household income were more likely to be supportive of the development of AI. A survey study in the Netherlands, using a representative panel of the Dutch population, studied trust in HCAI and found that the sociodemographic characteristics associated with higher levels of trust were being male, having a higher level of education, being employed or a student, and having not stayed in hospital in the past 12 months [11]. It is suggested that those who are less likely to suffer from the negative impacts of AI are more supportive of its implementation [10–12].

We conducted a survey to examine whether Australians’ attitudes toward HCAI vary with different sociodemographic characteristics.

## METHOD

Our aims for this study were threefold. We aimed to (1) examine the sociodemographic variables associated with support for AI in Australia, (2) examine the sociodemographic variables associated with support for HCAI, and (3) determine whether sociodemographic characteristics were associated with different preferences in AI-integrated healthcare.

This paper reports results from an analysis of the Australian Values and Attitudes toward AI (AVA-AI) survey. The survey was conducted with the Social Research Centre’s *Life In Australia* (LIA) study, which regularly engages a representative panel of Australians in independent surveys [13]. A shortened version of the AVA-AI questionnaire was included in the 36^th^ wave of the LIA study, disseminated in March 2020. The full version of the questionnaire was disseminated to a non-probabilistically sampled online panel. We used the shortened version of the questionnaire as a reference survey to produce weights for the non-probability sample that account for characteristics that influence people’s propensity to participate in the online panel. A more detailed description of the data collection and weighting methodology is provided in Isbanner et al. [14]. For this analysis, we report on results from the weighted non-probability sample using data obtained from the full questionnaire.

### Predictor variables

We selected predictor variables analogous to other surveys on public attitudes toward AI [10,15]. These variables included age group, gender, self-identification as having a chronic health condition or disability, living in a capital city, highest level of educational attainment, area socio-economic advantage (henceforth referred to as Socio-Economic Index for Areas [SEIFA])^1^, household income, self-reported health, and speaking a language other than English at home.

We removed any responses where the participant had not responded to all predictor and outcome variables (N=17). One participant identified with a gender outside of the male/female binary. This response was removed [17], and the limitations of this will be discussed further below. N=1983 responses were analysed.

We calculated Spearman’s rho coefficients to identify multicollinearity between predictor variables (Table 1). Some pairs of variables were moderately correlated. Those with high self-reported health status were less likely to identify as having a disability, and those living in a capital city were more likely to live in postcodes with less socioeconomic disadvantage. We deemed these moderate correlations unlikely to have a detrimental effect on model fitting or interpretation.

**Table 1.** correlation matrix of predictor variables (Spearman’s Rho coefficients). 0 indicates no correlation. Coefficients closer to 1 or -1 indicate stronger positive and negative correlation, respectively.

|  |  |  |  |  |  |  |  |  |  |
| --- | --- | --- | --- | --- | --- | --- | --- | --- | --- |
| Self-reported disability | 0.04 |  |  |  |  |  |  |  |  |
| Age group | -0.16 | 0.23 |  |  |  |  |  |  |  |
| Education | 0.26 | -0.12 | -0.22 |  |  |  |  |  |  |
| Gender | 0.13 | 0.04 | 0.11 | 0.05 |  |  |  |  |  |
| Household income | 0.07 | -0.22 | -0.17 | 0.24 | 0.04 |  |  |  |  |
| Speaks languages other than English at home | 0.20 | -0.13 | -0.23 | 0.24 | 0.00 | 0.02 |  |  |  |
| Living in a capital city | 0.08 | -0.14 | -0.19 | 0.18 | -0.01 | 0.12 | 0.22 |  |  |
| SEIFA | 0.05 | -0.14 | -0.06 | 0.15 | -0.01 | 0.11 | 0.08 | 0.32 |  |
| Self-reported health status | 0.08 | -0.34 | -0.23 | 0.14 | -0.02 | 0.17 | 0.10 | 0.08 | 0.10 |
|  | computer science experience | Self-reported disability | Age group | Education | Gender | Household income | languages other than English | Living in a capital city | SEIFA index |

### Outcome variables

Eleven outcome variables were selected for the three aims of the study (Table 2). Item 1 replicated a question from Zhang and DaFoe’s study [10], asking participants to indicate their level of support for the development of AI on a five-point semantic scale from *strongly oppose* to *strongly support*. Item 2 was a question that asked participants to consider their support for HCAI in a scenario where an unexplainable algorithm was being used to analyse patient health records and suggest treatments. Item 3 asked participants to consider their support for an algorithm that diagnosed diseases more accurately than physicians but required patients to share their health record. Item 5 asked participants to consider their support for HCAI in a scenario where its development leads to physicians becoming less skilled at tasks that were replaced by AI. Each of these questions asked participants to indicate their level of support on a five-point scale.

**Table 2.** Aims and outcome variables.

| Aim | Items used | Predictor variables |
| --- | --- | --- |
| Aim 1: examine the sociodemographic variables associated with support for AI in Australia | 1. Level of support for the development of AI | • Gender<br>• Age<br>• Self-identifies as having a chronic health condition or disability<br>• Education<br>• Household income<br>• Speaks languages other than English at home<br>• Resides in a capital city<br>• SEIFA<br>• Self-reported health |
| Aim 2: examine the sociodemographic variables associated with support for HCAI | 2. Level of support for HCAI that is unexplainable |  |
|  | 3. Level of support for HCAI that requires sharing personal data |  |
|  | 4. Level of support for HCAI that leads to clinician deskilling |  |
| Aim 3: determine whether sociodemographic characteristics were associated with different preferences in AI-integrated healthcare | 5. Importance of explainability |  |
|  | 6. Importance of getting an answer quickly |  |
|  | 7. Importance of getting an accurate answer |  |
|  | 8. Importance of being able to talk to a person about one's health |  |
|  | 9. Importance of knowing who is responsible for one's care |  |
|  | 10. Importance of reducing health system costs |  |
|  | 11. Importance of knowing the system treats everyone fairly |  |

Items 5-11 were preceded by a scenario asking participants to imagine a situation where an algorithm was reading a medical test, diagnosing them with a disease, and recommending treatments. Participants were asked to consider the importance of (5) explainability, (6) speed, (7) accuracy, (8) human oversight, (9) accountability, (10) cost to the healthcare system, and (11) equity. Participants responded on a five-point scale from *not at all important* to *very important*. Each outcome variable was recoded to binary categories, where the two highest categories (i.e., strongly support & somewhat support, very important & extremely important) were recoded to 1 and remaining categories were coded to 0.

### Statistical analysis

We generated frequency tables that incorporated the survey weights using the *questionr* package [18].We fit separate multiple logistic regression models for each of the outcome variables, using the same suite of sociodemographic variables as predictors for each. All analyses were conducted in R [19]. The *survey* package [20] was used to incorporate survey weights in the analysis and calculation of standard errors. Odds ratios (ORs) are reported with accompanying P-values and 95% confidence intervals. We considered results significant where *P* < 0.05 and commented on all results where *P* < 0.10.

## RESULTS

N=1983 responses were analysed. Weighted and unweighted sample demographics are shown in Table 3. Weights primarily affected distributions in self-reported health, chronic health condition or disability status, and speaking languages other than English at home.

**Table 3.** Weighted and unweighted sample demographics.

|  |  | Unweighted |  | Weighted |  |
| --- | --- | --- | --- | --- | --- |
|  |  | N | % | N | % |
| Computer science or programming experience | No | 1598 | 85.0% | 1603.4 | 85.3% |
|  | Yes | 281 | 15.0% | 275.6 | 14.7% |
| Has chronic health condition or disability | No | 1361 | 72.4% | 1457.2 | 77.6% |
|  | Yes | 518 | 27.6% | 421.8 | 22.4% |
| Age group | 18-34 | 572 | 30.4% | 593.2 | 31.6% |
|  | 35-54 | 630 | 33.5% | 640.5 | 34.1% |
|  | 55+ | 677 | 36.0% | 645.3 | 34.3% |
| Highest level of educational attainment | High School | 603 | 32.1% | 632.3 | 33.7% |
|  | Trade cert/Diploma | 630 | 33.5% | 709.3 | 37.7% |
|  | Bachelor's degree | 452 | 24.1% | 374.7 | 19.9% |
|  | Postgraduate Degree | 194 | 10.3% | 162.7 | 8.7% |
| Gender | Female | 947 | 50.4% | 968.1 | 51.5% |
|  | Male | 932 | 49.6% | 910.9 | 48.5% |
| Household income | <\$500 pw | 361 | 19.2% | 340.2 | 18.1% |
| | \$500 - \$1999 pw | 1095 | 58.3% | 1051.5 | 56.0% |
| | \$2000+ pw | 423 | 22.5% | 487.3 | 25.9% |
| Speaks languages other than English at home | No | 1598 | 85.0% | 1473.8 | 78.4% |
|  | Yes | 281 | 15.0% | 405.2 | 21.6% |
| Lives in capital city | No | 626 | 33.3% | 626.7 | 33.4% |
|  | Yes | 1253 | 66.7% | 1252.3 | 66.6% |
| SEIFA score | Most disadvantage | 281 | 15.0% | 294.9 | 15.7% |
|  | Moderate | 1185 | 63.1% | 1160.2 | 61.7% |
|  | Least disadvantage | 413 | 22.0% | 423.8 | 22.6% |
| Self-reported health | Excellent/very good | 735 | 39.1% | 1015.1 | 54.0% |
|  | Good/fair/poor | 1144 | 60.9% | 863.9 | 46.0% |

### Support for development of AI

Logistic regression results are displayed in Figure 1 with weighted proportions in Supplementary File 1. Overall, 56.7% of the weighted sample supported the development of AI. Support was significantly higher amongst those with computer science experience (weighted proportion supportive=72.1%; OR=1.89; *P=*0.001) compared to those without such experience; those with moderate (55.6%; OR=1.39; *P=*0.043) or high (66.3%; OR=1.90; *P*= 0.002) household incomes compared to those with low income; and those with trade certificates/diplomas (57.4%; OR=1.37; *P*=0.028), Bachelor’s degrees (65.6%; OR=1.61; *P=*0.008), and postgraduate degrees (69.0%; OR=1.75; *P=*0.022), compared to those with only high school-level education.

**Figure 1.**
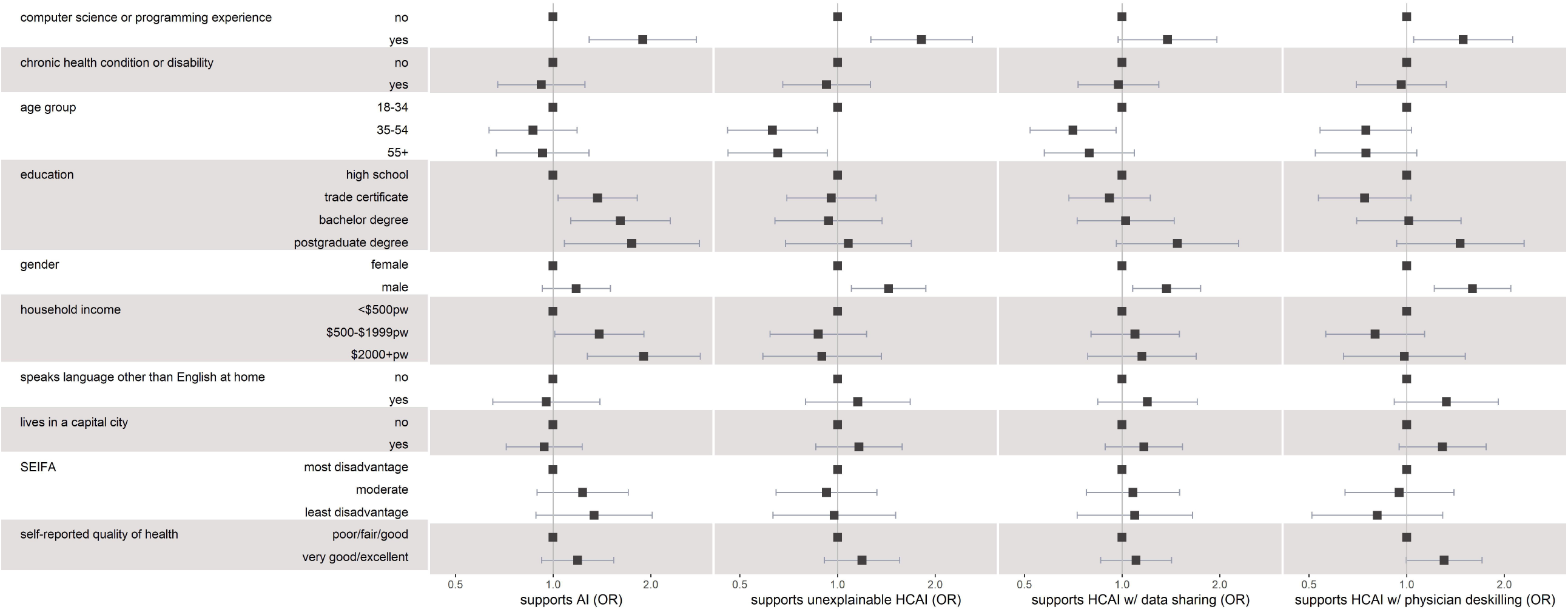
Odds ratio plot of weighted logistic regression results. Error bar indicates 95% confidence interval. Index categories displayed with OR=1. Plots indicate (1) participants’ level of support for artificial intelligence, (2) participants’ level of support for unexplainable AI in healthcare, (3) participants’support for AI in healthcare that necessitates sharing data, and (4) participants support for HCAI that leads to physician deskilling.

### Support for the development for HCAI and trade-offs

Participants were asked to consider whether they supported the development of HCAI in three scenarios. Across the weighted sample, only 27.0% were supportive of HCAI that led to physician deskilling, 28.7% were supportive of unexplainable HCAI, and 41.9% were supportive of HCAI that necessitated sharing personal data. Logistic regression results are displayed in Figure 1.

Support for unexplainable HCAI was significantly higher amongst those with computer science experience (43.4%; OR=1.82; *P=*0.001) and males (32.5%; OR=1.44; *P=*0.007). Support was significantly lower amongst those aged 35–54 (25.3%; OR=0.63; *P=*0.005) and those aged 55+ (25.0%; OR=0.65; *P*=0.018) compared to those aged 18–34 (36.4%).

Support for AI that necessitates data sharing was significantly higher amongst males (46.0%; OR=1.37; *P*=0.011). Those with a postgraduate degree (55.5%; OR=1.48; *P*=0.076) and computer science experience (53.1%; OR=1.38; *P*=0.071) also appeared to be more supportive of HCAI that requires sharing data. Participants aged 35–54 (38.4%; OR=0.71; P=0.025) were less likely than those aged 18–34 (48.4%) to be supportive of HCAI that necessitates data sharing.

Support for HCAI that leads to physician deskilling was significantly higher amongst those with computer science experience (40.0%; OR=1.49; *P*=0.025) and males (31.6%; OR=1.60; *P*=0.001). Support was lower, although this was not statistically significant, for those aged 35–54 (25.5%; OR=0.75; *P*=0.081), those with trade certificates or diplomas (21.6%; OR=0.74; *P*=0.076) compared to those with high school level education, and those with lower self-reported health (23.1%; OR=0.77; *P*=0.053).

The analysis did not show an association between household income, living in areas with less social disadvantage, living in a capital city, speaking languages other than English at home, or having a chronic health condition/disability and support for the HCAI trade-offs.

### Importance of different features in AI-integrated healthcare

Participants were asked to respond to a series of questions about the importance of various aspects of HCAI implementation. Logistic regression results can be found in Figure 2 and weighted proportions for each subgroup can be found in Supplementary File 2. Across all sociodemographic groups, accuracy was the feature most regarded as important, and reducing costs to the healthcare system was least likely to be regarded as important followed by speed.

**Figure 2.**
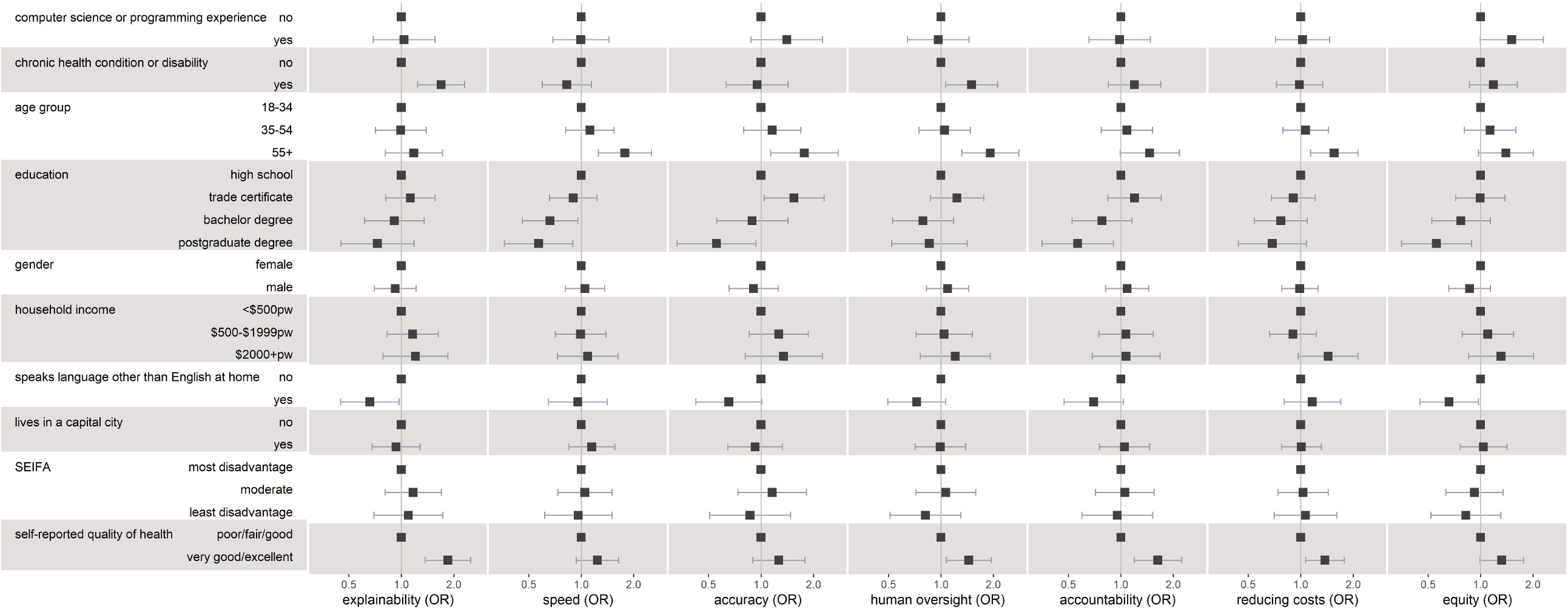
Odds ratio plot of weighted logistic regression results. Error bar indicates 95% confidence interval. Index categories displayed with OR=1. Plots indicate level of importance attributed to each aspect of AI-enabled care.

#### Socioeconomic characteristics

Socioeconomic factors had little effect on perceived importance of the features. Having a high (>ä2000pw) income had a weak positive effect on perceived importance of reducing costs to the healthcare system (64.5%; OR=1.44; *P*=0.073). SEIFA was not associated with perceived importance for any of the features.

#### Demographic characteristics

Demographic characteristics had some associations with perceived importance of the features. Those who spoke languages other than English at home were significantly less likely to regard explainability (68.0%; OR=0.66; *P*=0.035) and equity (65.1%; OR=0.66; *P*=0.035) as very/extremely important. They were also perhaps less likely to perceive accuracy (77.7%; OR=0.65; *P*=0.056) and accountability (70.6%; OR=0.70; *P*=0.074) as very/extremely important. Those aged over 55 were more likely than those aged 18–34 to perceive all features as very important, particularly human oversight (85.0%; OR=1.92; *P*=0.001), however this effect was not significant for equity and explainability. Gender and living in a capital city had no significant association with any of the features.

#### Educational characteristics

Those with postgraduate degrees were less likely than those with a high school-level education to see accuracy (73.9%; OR=0.55; *P*=0.027) equity (64.0%; OR=0.56; *P*=0.014), speed (61.1%; OR=0.57; *P*=0.015), and accountability (61.1%; OR=0.57; *P*=0.018) as very/extremely important. Those with computer science or programming experience were slightly more likely to see equity (76.0%; OR=1.51; *P*=0.052) as very/extremely important.

#### Health-related characteristics

Those with higher self-reported health were significantly more likely to perceive all features as important, except for equity (at *P* = 0.056), speed, and accuracy. Those who identified as having a chronic health condition were significantly more likely than those who did not to perceive explainability (81.1%; OR=1.69; *P*=0.001) and human oversight (83.2%; OR=1.5; *P*=0.02) as very/extremely important.

## DISCUSSION

In this study we examined sociodemographic differences in preference for healthcare AI using a large weighted Australian sample that was calibrated to the LIA probability sample using a range of behavioural and lifestyle questions, as well as major socio-demographic variables. Overall, 56.7% (95%CI=53.8%-59.0%) of the participants were supportive of the development of AI, slightly lower than results from another recent Australian study that also used an online panel, which found 62.4% were supportive [15]. In a separate analysis of the same AVA-AI survey, combining the LIA probability sample results with the online panel results [14], it was found that 60.3% (95%CI=58.4%-62.0%) of Australians were supportive of the development of AI. In the unweighted non-probability sample, 54.8% (95%CI=52.5%-57%) of participants supported the development of AI, suggesting that the use of an extensive set of variables in the weighting led to some improvement, but the potential of self-selection in online panels may not have been corrected fully by the sophisticated weighting methodology.

Similar to Zhang and DaFoe’s [10] study in the US, we found that support for the development of AI was higher among those with computer science experience, higher levels of education, and higher household incomes. It has been suggested that support for AI is lower amongst groups with less education and more social disadvantage, whose livelihoods may be more threatened by automation [10,12]. The potential for AI to threaten people’s livelihoods through taking jobs appears to be a poignant concern in Australia, where Selwyn et al.[15] found that the prospect of automation and job loss was the most commonly mentioned fear amongst their Australian sample. Results from our survey appear to support these findings, where metrics for social advantage (i.e. household income and education) were strongly associated with support for development of AI.

The sociodemographic characteristics associated with support for HCAI were different to those associated with support for AI in general. The items assessing support for HCAI required participants to consider whether they supported the development of HCAI, on balance, when it involved a trade-off (lack of explainability, data sharing, or physician deskilling). For each of these questions, household income and education were no longer strong predictors of support, and in some cases had a slight negative effect on support. Our findings do not appear to link socioeconomic advantage or disadvantage to attitudes about HCAI.

The characteristics that we found to be consistent predictors of support for HCAI and their specified tradeoffs were having computer science experience, being male, and being aged 18–34. These three characteristics are similar to those Selwyn et al. [15] found were associated with higher levels of familiarity with AI. This suggests that groups with more perceived familiarity with AI may be also more tolerant of its potential risks.

Our investigation into subgroup differences in the perceived importance of features of AI-integrated healthcare found that accuracy was regarded as particularly important by all subgroups. This differs from Ploug et al [21] who found, in a choice experiment in Denmark, that factors like explainability, equity, and physicians being responsible for decisions were regarded as more important than accuracy. The Danish experiment, however, offered the qualifier that the algorithm would at least be as accurate as a human doctor, whereas our questionnaire did not. Further research could test whether algorithmic performance is more important than other features in circumstances where there are no assurances that the algorithm is as good as a human doctor.

Health-related characteristics such as self-reported health and having a chronic health condition or disability had a particularly strong effect on perceived importance, where more importance was attributed to traditionally human aspects of healthcare like explainability, human oversight, and accountability. Subgroups that were more likely to be supportive of HCAI were not necessarily more likely to see the features of care that they were trading off as less important. Whilst those who identified as male, those aged 18–34, and those with computer science or programming experience were more likely to support the development of unexplainable AI in healthcare, they were just as likely as others to perceive explainability (“*knowing why a decision is made”*) as a very/extremely important aspect of AI-integrated care. This hints at a complex relationship between people’s support for the development of HCAI and their willingness to make compromises to their healthcare.

### Limitations

Given the quickly shifting landscape around AI, it is possible that public support for AI has changed in the two years since the questionnaire was administered. In addition, the AVA-AI survey includes an online panel obtained by non-probability sampling, which is subject to self-selection biases. The weighting methodology assists in reducing these effects by accounting for more than basic demographic variables, such as age by education, gender, household structure, language spoken at home, self-reported health, early adopter status, and TV streaming. Any selection effects due to the prediction variables included in the analysis are also accounted for. However, it is possible that support for HCAI is mediated by confounding factors not considered in the weighting methodology or included in the analysis.

One key population that were not represented in the study were those who identified as a gender outside of the male/female binary. Only one participant identified as a gender outside of the binary, and they were excluded from the analysis due to insufficient participant numbers to form a third gender category. Given that support for AI is lower amongst certain marginalised groups, consulting gender diverse individuals about their support for AI is an important consideration for future research.

Finally, the present study is a cross-sectional analysis which cannot infer causation between any of the predictor and outcome variables. Whilst we found an association between certain sociodemographic characteristics such as education, and outcomes such as level of support for AI, we cannot ascertain the reasons for this association. These reasons are likely complex and multi-faceted and should be explored in further research.

## CONCLUSION

Respondents who reported having greater ill-health or disability were more likely to consider human aspects of healthcare, such as explainability, human oversight, and accountability, as important. Whilst factors indicating socioeconomic advantage (higher income, higher education) were associated with general support for AI, these factors were not necessarily related to support for HCAI scenarios. Instead, support for HCAI scenarios was notably higher amongst males, younger people, and those with computer science or programming experience. Based on other research, these groups may have a higher level of familiarity with AI. Further research should examine the relationship between familiarity with AI and support for the development of AI.

^1^ This study used the Australian Bureau of Statistics’ Socioeconomic Indexes for Areas (SEIFA) to measure the relative advantage and disadvantage of areas [16]. Participants were classified into quintiles based on the SEIFA of their area (i.e.postcode) of residence, with those in quintiles 4 and 5 coded as ‘ least socioeconomic disadvantage’, those in quintiles 2 and 3 coded ‘ moderate disadvantage’, and those in quintile 1 coded as ‘ most socioeconomic disadvantage’.

## Data Availability

Due to ethical restrictions, data for this manuscript are not available.

## ACKNOWLEDGEMENTS

Statistical consulting was provided by Dr Brad Wakefield at the National Institute for Applied Statistics Research Australia (NIASRA) Statistical Consulting Centre.

## CONTRIBUTERSHIP STATEMENT

EF contributed to the design of the study, cleaned the data, conducted the analysis, and drafted the manuscript. PO contributed to the design of the survey instrument, contributed to the design of the study, oversaw the analysis, and edited the manuscript. DS contributed to the design of the survey and edited the manuscript. ABM and YA edited the manuscript. SC contributed to the design of the survey instrument, contributed to the design of the study, and edited the manuscript. All authors approved the final version of the manuscript for publication.

