## Supplementary File 1 for "Measures of socioeconomic advantage are not independent predictors of support for healthcare AI: subgroup analysis of a national Australian survey"

### Supplementary file 1 -. Weighted proportions of the sample who supported the development of AI (aim 1) and applications of HCAI (aim 2).

| | | Support the development of AI | Support for unexplainable AI in healthcare | Support for HCAI that necessitates data sharing | Support for HCAI that leads to physician deskilling |
|---|---|---|---|---|---|
| | ALL | 56.7% | 28.7% | 41.9% | 27.0% |
| Has computer science or programming experience | No | 54.0% | 26.1% | 39.9% | 24.8% |
| | Yes | 72.1% | 43.4% | 53.1% | 40.0% |
| Self-identifies as having a chronic illness or disability | No | 58.1% | 29.4% | 42.7% | 27.9% |
| | Yes | 51.6% | 26.2% | 39.1% | 24.1% |
| Age group | 18-34 | 61.2% | 36.4% | 48.4% | 32.9% |
| | 35-54 | 55.3% | 25.3% | 38.4% | 25.5% |
| | 55+ | 53.8% | 25.0% | 39.3% | 23.0% |
| Education | high school ed | 47.4% | 26.7% | 39.4% | 25.8% |
| | Diploma or trade cert | 57.4% | 26.9% | 38.7% | 21.6% |
| | Bachelor degree | 65.6% | 32.1% | 46.2% | 32.7% |
| | Postgraduate degree | 69.0% | 35.9% | 55.5% | 41.9% |
| Speaks languages other than English at home | No | 55.6% | 26.9% | 39.9% | 24.4% |
| | Yes | 60.4% | 35.3% | 49.1% | 36.4% |
| Gender | Female | 53.9% | 25.1% | 38.0% | 22.6% |
| | Male | 59.6% | 32.5% | 46.0% | 31.6% |
| Household income | <$500 pw | 45.9% | 29.7% | 38.6% | 28.2% |
| | $500 - $1999 pw | 55.6% | 27.5% | 41.4% | 24.5% |
| | $2000+ pw | 66.3% | 30.5% | 45.2% | 31.6% |
| Region | Outside of capital city | 54.3% | 24.8% | 37.2% | 22.1% |
| | Capital City | 57.8% | 30.6% | 44.2% | 29.5% |
| SEIFA | most disadvantage | 49.9% | 27.6% | 38.1% | 25.8% |
| | moderate | 56.5% | 27.9% | 41.5% | 26.8% |
| | least disadvantage | 61.8% | 31.7% | 45.5% | 28.3% |
| Self reported health | good/fair/poor | 52.4% | 25.6% | 39.3% | 23.1% |
| | excellent/very good | 60.3% | 31.3% | 44.1% | 30.4% |
