## Supplementary File 2 for "Measures of socioeconomic advantage are not independent predictors of support for healthcare AI: subgroup analysis of a national Australian survey"

**Supplementary file 2 - -. Weighted proportions of the sample who felt each aspect of AI-integrated care was very/extremely important.**

| | | Explainability | Speed | Accuracy | Human oversight | Accountability | Reducing costs | Equity |
|---|---|---|---|---|---|---|---|---|
| | ALL | 76.0% | 70.3% | 85.0% | 77.1% | 78.4% | 57.9% | 73.5% |
| Has computer science or programming experience | No | 76.2% | 70.9% | 85.0% | 77.6% | 78.9% | 58.0% | 73.1% |
| | Yes | 75.1% | 66.4% | 84.7% | 74.2% | 75.2% | 57.0% | 76.0% |
| Self-identifies as having a chronic illness or disability | No | 74.6% | 70.8% | 84.7% | 75.3% | 77.7% | 58.5% | 72.6% |
| | Yes | 81.1% | 68.3% | 85.9% | 83.2% | 80.6% | 55.9% | 76.8% |
| Age group | 18-34 | 74.4% | 65.6% | 81.4% | 71.7% | 74.9% | 54.9% | 69.9% |
| | 35-54 | 74.5% | 67.9% | 83.7% | 74.2% | 76.7% | 55.9% | 72.8% |
| | 55+ | 79.0% | 77.0% | 89.4% | 85.0% | 83.1% | 62.5% | 77.6% |
| Education | high school ed | 76.7% | 74.2% | 84.9% | 78.4% | 79.9% | 60.0% | 75.9% |
| | Diploma or trade cert | 78.7% | 71.8% | 89.5% | 81.1% | 82.3% | 58.0% | 75.9% |
| | Bachelor degree | 72.8% | 64.8% | 81.3% | 69.7% | 73.2% | 55.9% | 69.4% |
| | Postgraduate degree | 69.2% | 61.1% | 73.9% | 71.7% | 67.1% | 53.5% | 64.0% |
| Speaks languages other than English at home | No | 78.2% | 71.3% | 87.0% | 79.6% | 80.5% | 57.7% | 75.9% |
| | Yes | 68.0% | 66.4% | 77.7% | 68.1% | 70.6% | 58.6% | 65.1% |
| Gender | Female | 76.6% | 69.4% | 85.1% | 75.7% | 77.5% | 57.6% | 74.3% |
| | Male | 75.5% | 71.2% | 84.8% | 78.6% | 79.2% | 58.2% | 72.8% |
| Household income | <$500 pw | 74.4% | 70.4% | 82.7% | 76.9% | 77.8% | 57.8% | 72.4% |
| | $500 - $1999 pw | 76.6% | 70.4% | 86.0% | 77.4% | 79.1% | 54.8% | 73.5% |
| | $2000+ pw | 75.9% | 70.0% | 84.4% | 76.5% | 77.1% | 64.5% | 74.4% |
| Region | Outside of capital city | 78.7% | 70.4% | 87.9% | 80.5% | 80.4% | 57.6% | 75.7% |
| | Capital City | 74.7% | 70.2% | 83.5% | 75.4% | 77.3% | 58.0% | 72.5% |
| SEIFA | most disadvantage | 74.9% | 70.4% | 85.4% | 78.9% | 79.0% | 56.8% | 76.2% |
| | moderate | 77.1% | 70.7% | 86.3% | 78.4% | 79.1% | 57.6% | 73.9% |
| | least disadvantage | 73.9% | 69.1% | 81.1% | 72.2% | 75.9% | 59.5% | 70.6% |
| Self reported health | good/fair/poor | 72.0% | 68.7% | 84.3% | 76.0% | 75.6% | 54.0% | 71.9% |
| | excellent/very good | 79.5% | 71.7% | 85.5% | 78.0% | 80.7% | 61.2% | 74.9% |
